# Impact of Clinical Trial Virtualization on Recruitment in Underserved Communities for a Type 2 Diabetes mHealth Intervention

**DOI:** 10.1101/2025.10.27.25338889

**Authors:** Elizabeth Campbell, Andrea Cassells, TJ Lin, Jacqeline Cortez Lainez, Arlene Smaldone, Pooja Desai, Haomiao Jia, George Hripcsak, Jonathan Tobin, Lena Mamykina

## Abstract

**Objective:** Approaches to randomized clinical trial (RCT) implementation greatly changed during the COVID-19 pandemic as clinical trials transitioned to largely virtual implementation; virtual RCT implementation may continue in the future. Without careful consideration, virtual RCTs may exacerbate the historical under-representation of women, the elderly, and racial and ethnic minorities that has occurred in past clinical trials. The following study presents an approach to virtualizing an RCT for an mHealth intervention.

**Materials and Methods:** Participants for the trial were recruited from Federally Qualified Health Centers in medically underserved areas of the New York City metropolitan area. Recruitment began in January 2020 but was paused in March 2020 due to the COVID-19 pandemic’s onset. The research team developed a virtual protocol for recruitment, onboarding, and study implementation.

**Results:** Study participants were predominantly from immigrant, low-income, and racial and ethnic minority groups. Our virtualization approach included an easier-to-understand consent form, expanded virtual training materials, and greater one-on-one attention during training. Our approach did not lead to significant changes in the population we recruited into the study or introduce additional biases and restriction.

**Discussion:** Our study represents an early investigation into how the change from in-person clinical trial recruitment and study implementation to virtual during the COVID-19 pandemic may impact recruitment of participants from medically underserved communities into RCTs.

**Conclusion:** This approach may be used in future trials testing mHealth and other technological interventions without exacerbating the under-representation of medically underserved populations.

**Highlights:**

- The COVID-19 pandemic accelerated the shift to virtual randomized clinical trials (RCTs), raising concerns about exacerbating the under-representation of marginalized groups.
- This study implemented a fully virtual RCT for a mHealth intervention with consideration of inclusivity.
- Participants were recruited from Federally Qualified Health Centers in medically underserved areas of New York City, with most coming from immigrant, low-income, and minority communities.
- Adaptations such as simplified consent, expanded virtual training, and individualized support ensured accessibility without altering participant demographics or introducing bias, providing a replicable model for equitable virtual trials.

## INTRODUCTION

Artificial intelligence (AI) is revolutionizing medical research and clinical care, through its impact on screening, disease detection, symptom monitoring, disease prediction and treatment.[1,2] As a result, medical interventions using AI, including mHealth interventions, are being increasingly investigated in randomized clinical trials (RCTs) so that these applications may be implemented at scale. [3–5]

However, RCTs often fall short in representing diverse patient populations, including women, racial and ethnic minorities, and older adults.[6–8] mHealth interventions often present high demands on technical and digital literacy, which may further exacerbate these disparities and introduce new barriers to participation in RCTs and disease self-management. [9] When clinical trials fail to be broadly representative of a population’s diversity, interventions may not translate well in real-world applications and can become differentially effective for vulnerable subgroups.[10,11] Furthermore, this lack of diversity creates numerous other downstream effects in clinical research such as under-representation in datasets and bias in machine learning models developed using these data.[12–14]

The COVID-19 pandemic adversely affected clinical research in numerous ways. Studies were delayed, placed on hold, and became inaccessible to patients and trial personnel for in-person study visits and follow-up.[15] As a result, recruitment, study visits, and other study implementation activities were changed from in-person to virtual by employing teleconferencing services to ensure the safety of participants, research and clinical practice staff.[16,17] This move toward digitalization has highlighted both the desirability and the viability of decentralized clinical trials utilizing technology and, if adopted, may reduce barriers to participation in RCTs even after the pandemic.[15]

In the future, RCTs may have a greater reliance on remote patient recruitment, onboarding, intervention, assessment, and follow-up monitoring. Remote patient recruitment and onboarding reduces geographic constraints so participants can be recruited from more locations; allowing patients to engage remotely may make trial participation easier for many.[18–20] However, a greater reliance on electronic communication devices, mobile apps, and other technologies in clinical trials may unintentionally exclude those of lower income, older age, and less formal education and/or technical literacy. This may result in vulnerable patient sub-populations being under-recruited for clinical trials that largely rely on virtual implementation.[20]

In this study, we present an approach to virtualizing the T2 Coach study, an RCT for a Type 2 Diabetes (T2DM) mHealth intervention that shifted from in-person to virtualized implementation during the COVID-19 pandemic. Representation in clinical trials for T2DM interventions is especially critical given the disparities in access to healthcare services and education. Moreover, racial and ethnic minorities and low-income patients have a higher prevalence of T2DM and associated comorbidities.[21,22] In this work, we describe the clinical trial virtualized protocol, participant recruitment, and implementation for the T2 Coach study, and compare demographic, clinical, and digital literacy characteristics of participants recruited in-person pre-pandemic versus virtually after the pandemic’s onset. This study aims to address the following research questions:

1. Are there significant differences in the demographic and clinical characteristics between patients recruited in-person and virtually in the T2 Coach study?
2. Does mobile device proficiency differ between patients recruited in-person and virtually in the T2 Coach study?

## METHODS

The T2 Coach study is a two-arm RCT which tested the efficacy of T2 Coach, a personalized behavioral mHealth intervention for individuals with T2DM that employs a conversational agent to help users to set and work towards achieving personalized T2DM self-management goals.

### Setting

Participants for the trial were recruited from six Federally Qualified Health Centers (FQHCs) in medically underserved areas of New York City and the surrounding metropolitan area that are part of Clinical Directors Network’s primary care practice-based research network.

### Study Population

Participants met study inclusion criteria if they (a) were a patient of the FQHC for ≥ 6 months and had a diagnosis of T2DM (b) had HbA1c ≥ 8.0, (c) were age 18 to 65 years, (d) owned a smartphone, and (e) were proficient in either English or Spanish. Participants were excluded if they (a) were pregnant, (b) had a severe cognitive impairment (recorded in patient chart), (c) had other serious illnesses (e.g. cancer diagnosis with active treatment, advanced stage heart failure, dialysis, multiple sclerosis, advanced retinopathy, recorded in patient chart) (d) planned to leave the FHQC in the next 12 months, or e) had participated in the pilot study preceding T2 Coach.[23]

### Patient Recruitment and Clinical Trial Virtualization

Study recruitment began in January 2020. Twenty-one patients were recruited in person from two participating sites. During this in-person recruitment phase, clinical trial personnel approached patients in FQHC waiting areas to introduce the study and conduct the screening to then obtain consent for participation. The study team also received referrals from FQHC staff for patients who were likely a good fit for the trial. Prior to the pandemic, participants were recruited in-person and provided written consent for participation and received an incentive in cash.

In March 2020, the T2 Coach study team paused recruitment due to the COVID-19 pandemic, restrictions on in-person gatherings in New York City, and to ensure the safety of research and site staff, as well as the FQHC patients. During the spring and summer of 2020, the team developed a new set of protocols and strategies for implementing all components of the project virtually, including recruitment, onboarding, training, and all follow-up and retention activities, and secured IRB approvals for the new set of procedures. These protocols included:

- Secure remote access to electronic health record systems of the participating FQHCs and protocols for screening patients for eligibility
- Site-approved virtual recruitment strategies, including direct outreach to eligible participants
- Remote screening and oral consent
- Remote delivery of intervention materials
- Downloading intervention software on participants’ smartphones and remote training on use of intervention
- Participant follow-up
- Electronic gift card payment of incentives (reloadable debit cards)

By August 2020, all components of the intervention had been switched to virtual and participant recruitment was resumed at the two sites that initiated recruitment pre-pandemic, and at additional four sites. To identify eligible participants for recruitment, the T2 Coach team obtained secure remote access to electronic health records at participating sites and received a list of potentially eligible participants from the FQHCs (which included patients with HbA1c ≥ 8 in the last month, age, most recent visit date, diagnosis of T2DM). The team reached out to patients via phone, which often required several phone calls. As a part of clinical trial recruitment, potential participants’ ability to use Zoom was also assessed. After virtualization, we developed and implemented a simpler, more easily understandable oral consent form to streamline the informed consent process. This revised consent form reduced the amount of time required by participants to stay on the phone to complete the informed consent process and lessen the chances of patients hanging up or refusing to participate because the screening, consent and baseline assessment would take too long.

Converting the T2 Coach study protocol to virtual implementation necessitated the introduction of a new eligibility criterion – an ability to download, install, and use Zoom for virtual training. Anticipating that virtual recruitment, onboarding, and intervention training procedures may present barriers to the target population, the research team introduced an enhanced virtual training package that included online video tutorials explaining different features of the intervention, and additional opportunities to consult the research team for extra explanations and training, in addition to the main training session.

Prior to virtualization, study participants conducted a face-to-face session with a recruitment coordinator (RC) who played a T2 Coach instructional video on the RC’s computer. The RC demonstrated how to use the T2 Coach app with the participant’s phone and provided a T2DM educational brochure. The in-person training session lasted approximately 1 hour. After virtualization, study participants were mailed training session materials including an educational brochure, a PowerPoint printout version of the T2 Coach instructional video, and a training manual with instructions on how to join a meeting and share their screen using Zoom. The RC scheduled a Zoom session and sent a Zoom link for the training session. During the virtual training session, the RC provided instructions on how to use the T2 Coach app via Zoom using a manualized training script. During this training session, the participants were asked to share their screen with researchers to enable research staff to observe their usage of the intervention, assist with problems, and correct mistakes. The virtual training session lasted 1-2 hours.

Table 1 summarizes the protocols for the T2 Coach study’s recruitment, onboarding, training, follow up, and retention for its in-person and virtualized implementation.

**Table 1.**
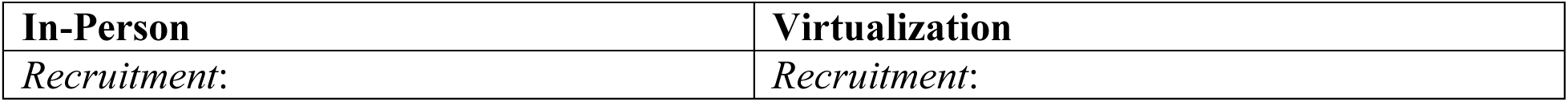
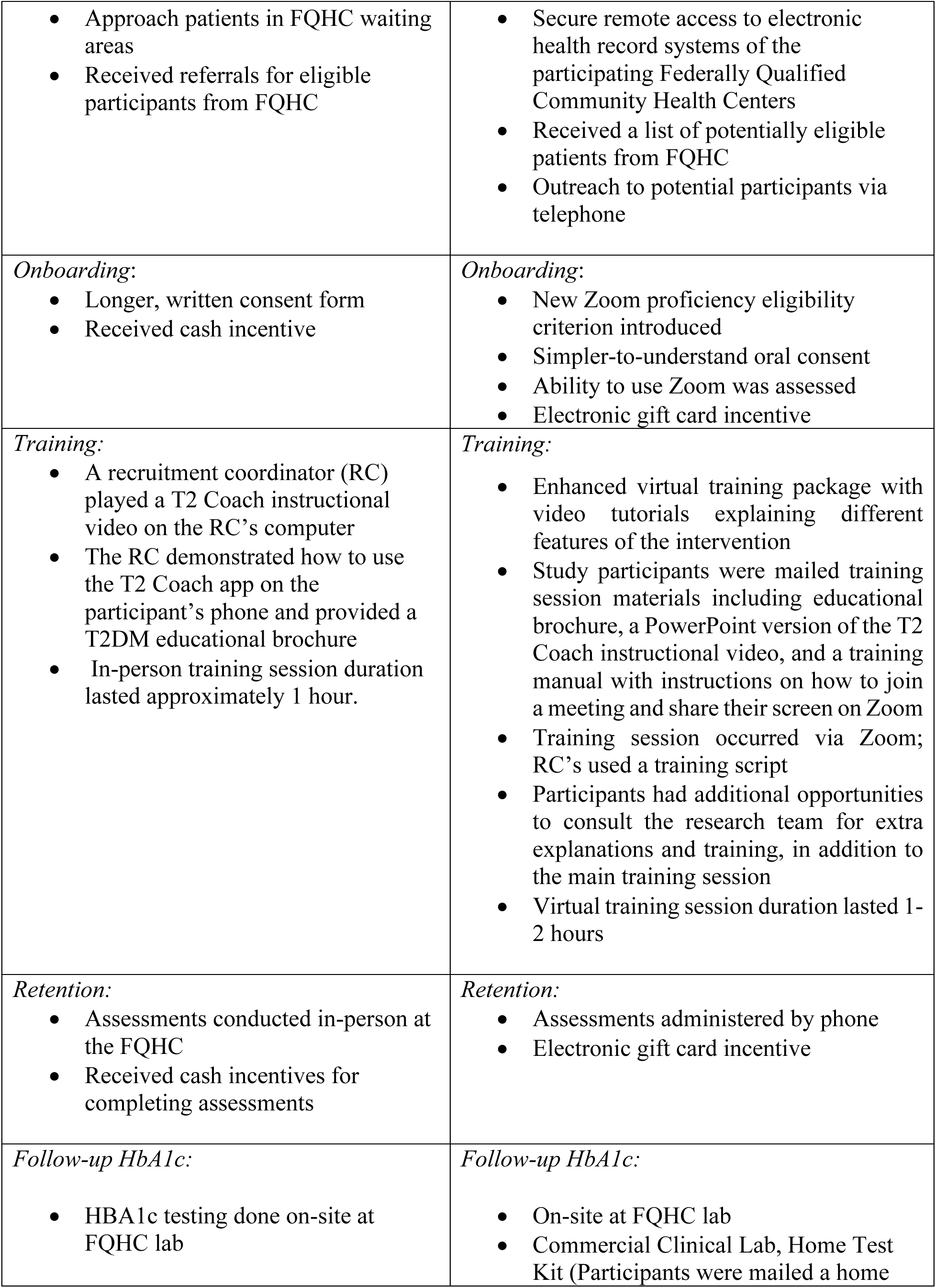
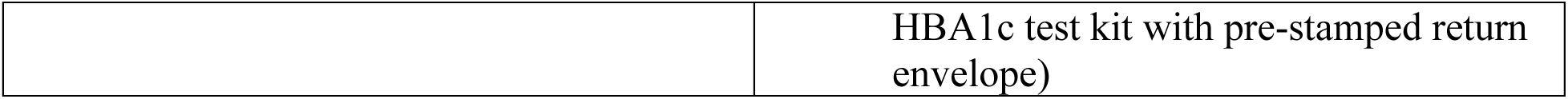
Comparison of In-Person with Virtualization Protocols for T2 Coach Study.

| **In-Person** | **Virtualization** |
| --- | --- |
| *Recruitment:* | *Recruitment:* |
| • Approach patients in FQHC waiting areas<br>• Received referrals for eligible participants from FQHC | • Secure remote access to electronic health record systems of the participating Federally Qualified Community Health Centers<br>• Received a list of potentially eligible patients from FQHC<br>• Outreach to potential participants via telephone |
| *Onboarding:*<br>• Longer, written consent form<br>• Received cash incentive | *Onboarding:*<br>• New Zoom proficiency eligibility criterion introduced<br>• Simpler-to-understand oral consent<br>• Ability to use Zoom was assessed<br>• Electronic gift card incentive |
| *Training:*<br>• A recruitment coordinator (RC) played a T2 Coach instructional video on the RC's computer<br>• The RC demonstrated how to use the T2 Coach app on the participant's phone and provided a T2DM educational brochure<br>• In-person training session duration lasted approximately 1 hour. | *Training:*<br>• Enhanced virtual training package with video tutorials explaining different features of the intervention<br>• Study participants were mailed training session materials including educational brochure, a PowerPoint version of the T2 Coach instructional video, and a training manual with instructions on how to join a meeting and share their screen on Zoom<br>• Training session occurred via Zoom; RC's used a training script<br>• Participants had additional opportunities to consult the research team for extra explanations and training, in addition to the main training session<br>• Virtual training session duration lasted 1-2 hours |
| *Retention:*<br>• Assessments conducted in-person at the FQHC<br>• Received cash incentives for completing assessments | *Retention:*<br>• Assessments administered by phone<br>• Electronic gift card incentive |
| *Follow-up HbA1c:*<br>• HbA1c testing done on-site at FQHC lab | *Follow-up HbA1c:*<br>• On-site at FQHC lab<br>• Commercial Clinical Lab, Home Test Kit (Participants were mailed a home |
|  | HBA1c test kit with pre-stamped return envelope) |

**Table 2.**
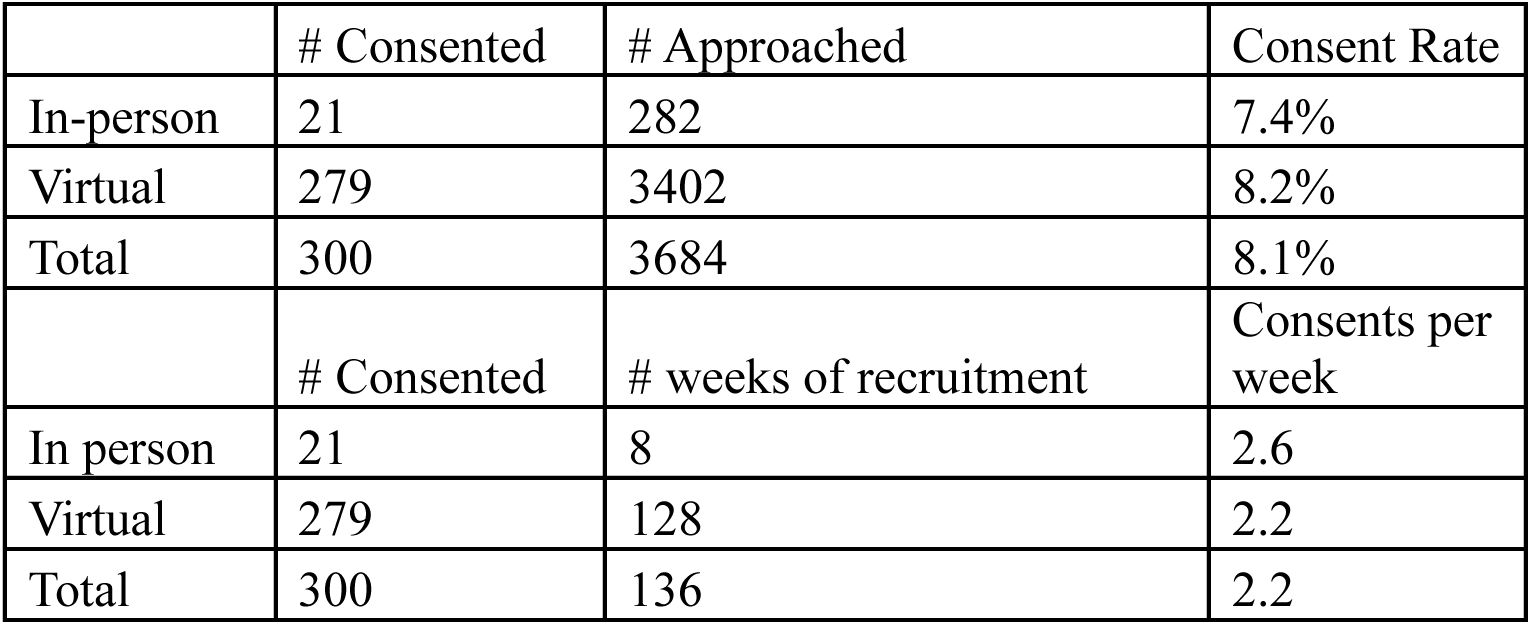
Consent rates and Consents per week for in-person and virtual recruitment efforts.

|  | # Consented | # Approached | Consent Rate |
| --- | --- | --- | --- |
| In-person | 21 | 282 | 7.4% |
| Virtual | 279 | 3402 | 8.2% |
| Total | 300 | 3684 | 8.1% |
|  | # Consented | # weeks of recruitment | Consents per week |
| In person | 21 | 8 | 2.6 |
| Virtual | 279 | 128 | 2.2 |
| Total | 300 | 136 | 2.2 |

### Baseline Data Collection

At the baseline visit, all subjects completed a demographic questionnaire, three questionnaires that assessed T2DM self-management and psychosocial measures, and a medical records review. The demographic questionnaire was an investigator-developed instrument that collected self-reported data on several items including age, race, ethnicity, gender, employment, income, and education. Current and past medical history and medications (including insulin therapy), along with HbA1c, fasting blood glucose, systolic and diastolic blood pressure, lipids, weight, and BMI was collected at baseline (additional measurements were taken at 6 months and 12 months but are not included in this paper).

During baseline enrollment, study participants also responded to a 16-question survey on self-perceived mobile device proficiency (MDPQ).[24] Participants self-described their proficiency for 16 questions about different mobile device activities, which were each considered as continuous variables in the data analysis. The 16 questions are grouped into 8 subscales: mobile device basics, communication, data and file storage, internet, calendar, entertainment, privacy, and trouble shooting and software management. The average score for the two questions that comprise a subscale is that participant’s subscale score ranging between 0-5. The eight subscale scores are summed for a final measure of total mobile device proficiency on a scale of 0-40.

### Randomization

Eligible participants (N=300) were randomized 1 to 1 to the intervention arm and the control arm. Randomization was stratified by site, language (English and Spanish), gender (male and female), and by those who were recruited in-person (n= 21) and those who were recruited virtually (n= 279). Randomization was in blocks of size=4. The randomization was done by a statistician who was blinded to the treatment assignments.

### Data Analysis

Descriptive statistics for the demographic and clinical variables collected at baseline were calculated to characterize the full study population. Patients were classified as Hispanic if their self-identified ethnicity was specified as Hispanic or Latino and were otherwise categorized by their response to the self-identified race question on the baseline questionnaire. The mean and standard deviation average response for each of the eight subscales and full score for the MDPQ was calculated for the total study population and then separately for the patient subgroups who were recruited in-person and virtually. The Shapiro-Wilk normality test was performed on the data. Because the normality assumption was rejected, the Wilcoxon signed-rank test was used to determine if there were statistically significant differences in mobile device proficiency measures as well as the demographic and clinical characteristics between the in-person and virtually recruited patient groups. The Shapiro-Wilk normality test revealed that data for three continuous variables (Age, HbA1c, and BMI at baseline) were normally distributed. The Wilcoxon-Mann-Whitney test was used to compare the continuous variables between the virtual and in-person subgroups, and the chi-squared test was used to compare education. All analyses were completed using R statistical software version 4.4.2.

## RESULTS

During in-person recruitment, 282 patients were approached and 21 consented for a consent rate of 7.4%. During virtual recruitment, 3,402 patients were approached and 279 consented, for a consent rate of 8.2%. In total, 3,684 patients were approached, and the consent rate was 8.1%. In-person recruitment occurred over the course of 8 weeks; there was an average of 2.6 consents obtained per week. Virtual recruitment occurred over the course of 128 weeks; there was an average of 2.2 consents per week. Three hundred consents occurred over the course of 136 weeks, leading to an average of 2.2 consents per week for the study.

Table 3 presents the clinical and demographic characteristics of the total baseline study population (n=300) and stratified by in-person (n=21) versus virtually recruited (n=279) participants. Over half of the study population was female (59.7%) and 61.0% of patients were Hispanic. Approximately one third of patients (30.7%) were Black or African American, and 63.7% were born in a country outside the US. More than half of participants had a High School degree or less (60.3%), were employed (61.3%), made a combined family income of less than $40,000 per year (81.9%), and spoke Spanish or a combination of Spanish and English at home (55.7%). Almost one third of patients (31.0%) did not have any insurance and 44.7% were enrolled in Medicaid. The mean participant age at baseline was 48.7 years, the mean HbA1c was 10.1, and the mean BMI was 33.3.

**Table 3.** Clinical and Demographic Characteristics of T2 Coach Study Population (N (%))

| Indicator | Full<br>Population<br>N = 300 | In-Person<br>N = 21 | Virtual<br>N= 279 | P-Value |
| --- | --- | --- | --- | --- |
| <u>*Gender*</u> |  |  |  |  |
| Male | 121 (40.3%) | 11 (52.4%) | 110 (39.4%) | 0.24 |
| Female | 179 (59.7%) | 10 (47.6%) | 169 (60.6%) |  |
| <u>*Race/Ethnicity*</u> |  |  |  |  |
| Hispanic | 183 (61.0%) | 6 (28.6%) | 177 (63.4%) | <0.01 |
| Asian | 8 (2.7%) | 1 (4.8%) | 7 (2.5%) |  |
| Black or African American | 92 (30.7%) | 12 (57.1%) | 80 (28.7%) |  |
| White | 11(3.7%) | 0 | 11 (3.9%) |  |
| Native Hawaiian or Other<br>Pacific Islander | 0 | 0 | 0 |  |
| American Indian/ Alaskan<br>Native | 1 (<1%) | 1 (4.8%) | 0 |  |
| Unknown | 0 | 0 | 0 |  |
| Multiple Race | 1 (<1%) | 0 | 1 (<1%) |  |
| Other | 4 (1.3%) | 1 (4.8%) | 3 (1.1%) |  |
| Refused | 0 | 0 | 0 |  |
| <u>*Insurance*</u> |  |  |  |  |
| Private | 36 (12.0%) | 3 (14.3%) | 33 (11.8%) | <0.01 |
| Medicare | 19 (6.3%) | 4 (19.0%) | 15 (5.4%) |  |
| Medicaid | 134 (44.7%) | 12 (57.1%) | 122 (43.7%) |  |
| Military/ VA | 0 | 0 | 0 |  |
| None | 93 (31.0%) | 0 | 93 (33.3%) |  |
| Other | 7 (2.3%) | 0 | 7 (2.5%) |  |
| Dual Enrollment (Medicare<br>and Medicaid) | 8 (2.7%) | 2 (9.5%) | 6 (2.2%) |  |
| Multiple (other) | 3 (1.0%) | 0 | 3 (1.1%) |  |
| <u>*Birth Location*</u> |  |  |  |  |
| United States/ Puerto Rico | 109 (36.3%) | 14 (66.7%) | 95 (34.1%) | <0.01 |
| Another Country | 191 (63.7%) | 7 (33.3%) | 184 (65.9%) |  |
| <u>*Language Spoken at Home*</u> |  |  |  |  |
| English | 122 (40.7%) | 14 (66.7%) | 108 (38.7%) | 0.11 |
| Spanish | 99 (33.0%) | 1 (4.8%) | 98 (35.1%) |  |
| Other | 4 (1.3%) | 1 (4.8%) | 3 (1.1%) |  |
| English and Spanish | 68 (22.7%) | 4 (19.0%) | 64 (22.9%) |  |
| English and Other | 5 (1.7%) | 1 (4.8%) | 4 (1.4%) |  |
| Spanish and Other | 2 (<1%) | 0 | 2 (<1%) |  |
| <u>Education Level</u> |  |  |  |  |
| Grades 1-8 | 53 (17.7%) | 1 (4.8%) | 52 (18.6%) | 0.19 |
| Grades 9-11 | 34 (11.3%) | 1 (4.8%) | 33 (11.8%) |  |
| High School Degree/GED | 94 (31.3%) | 8 (38.1%) | 86 (30.8%) |  |
| Some College but no Degree | 45 (15.0%) | 6 (28.6%) | 39 (14.0%) |  |
| Technical School Certificate | 18 (6.0%) | 3 (14.3%) | 15 (5.4%) |  |
| Associates Degree | 17 (5.7%) | 1 (4.8%) | 16 (5.7%) |  |
| Bachelor’s Degree | 29 (9.7%) | 1 (4.8%) | 28 (10.0%) |  |
| Graduate Degree | 10 (3.3%) | 0 | 10 (3.6%) |  |
| <u>Employment</u> |  |  |  |  |
| Yes | 184 (61.3%) | 10 (47.6%) | 174 (62.4%) | 0.18 |
| No | 116 (38.7%) | 11 (52.4%) | 105 (37.6%) |  |
| <u>Combined Family Income</u> |  |  |  |  |
| < \$10,000 | 103 (34.3%) | 6 (28.6%) | 97 (34.8%) | 0.11 |
| \$10,000- \$19,999 | 76 (25.3%) | 4 (19.0%) | 72 (25.8%) | |
| \$20,000- \$39,999 | 67 (22.3%) | 3 (14.3%) | 64 (22.9%) | |
| \$40,000- \$59,999 | 33 (11.0%) | 4 (19.0%) | 29 (10.4%) | |
| >\$60,000 | 15 (5.0%) | 1 (4.8%) | 14 (5.0%) | |
| Unknown | 6 (2.0%) | 3 (14.3%) | 3 (1.1%) |  |
| <u>Marital Status</u> |  |  |  |  |
| Married/Living with partner | 133 (44.3%) | 6 (28.6%) | 127 (45.5%) | 0.57 |
| Separated/ Divorced | 44 (14.7%) | 6 (28.6%) | 38 (13.6%) |  |
| Widowed | 14 (4.7%) | 3 (14.3%) | 11 (3.9%) |  |
| Never Married | 109 (35.8%) | 6 (28.6%) | 103 (36.9%) |  |
| Age in Years (Mean (SD)) | 48.7 (9.1) | 51 (7.8) | 48.5 (9.2) | 0.29 |
| HbA1c at Baseline (Mean (SD)) | 10.1 (1.7) | 10.3 (1.5) | 10.1 (1.7) | 0.37 |
| BMI (Mean (SD)) | 33.3 (7.7) | 35.7 (6.8) | 33.2 (7.7) | 0.04 |

When comparing the clinical and demographic characteristics of participants recruited virtually versus in-person, there were no statistically significant differences in gender, language spoken at home, education level, employment status, combined family income, marital status, age, and HbA1c at baseline. Statistically significant differences were observed in race/ethnicity, health insurance, birth location, and BMI. A much higher proportion of participants recruited virtually were Hispanic (63.4% versus 28.6%) and a higher proportion of participants recruited in-person were African American (57.1% versus 28.7%). One third of participants recruited virtually did not have health insurance (33.3%) while all participants recruited in-person had some type of insurance; Medicaid enrollment was higher for participants recruited in-person (57.1% versus 43.7%). Nearly two thirds of participants recruited in-person were born in the US or Puerto Rico, while the opposite was true for participants recruited virtually; 65.9% of virtually recruited participants were from another country. The mean BMI at baseline for participants recruited in-person was 35.7 versus 33.2 for patients recruited virtually.

Table 4 presents the analysis from the MDPQ administered to all study participants at baseline enrollment. MDPQ mean subscale scores ranged from 3.38 −4.76 for patients recruited in-person and ranged from 3.40-4.66 for patients recruited virtually. Participants from both groups both scored lowest in self-perceived proficiency in data file and storage measures and scored highest in performing basic actions with their mobile device (such as navigating on-screen menus, using the onscreen keyboard, etc.). No statistically significant differences were identified in mobile device proficiency between the two study groups for all eight measures (p< 0.05) and there were no significant differences in total MDPQ scores between the two study groups.

**Table 4.**
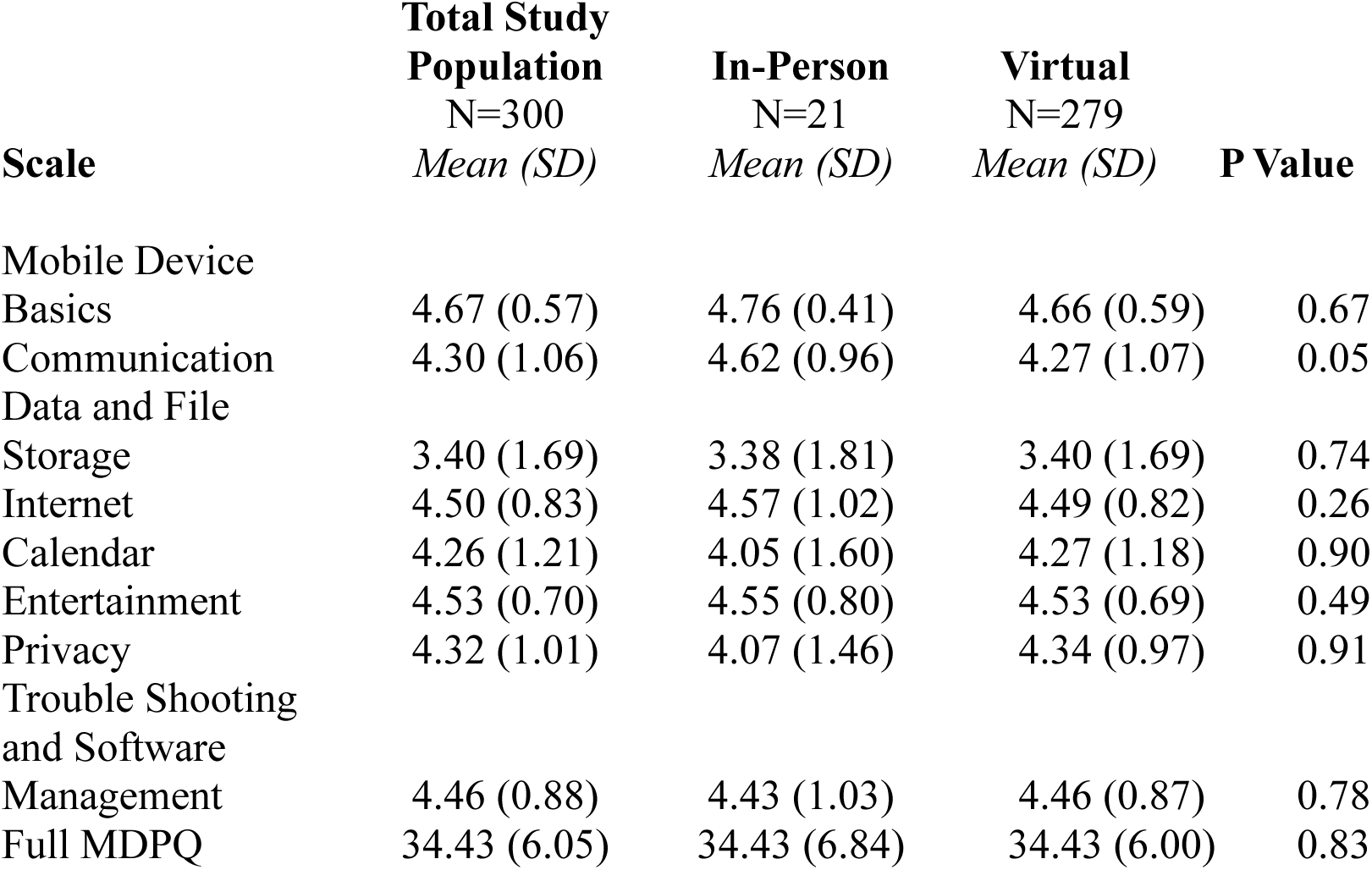
Mobile Device Proficiency Questionnaire Results for T2 Coach Study Population.

## DISCUSSION

Many clinical trials had to transition from primarily in-person to a largely virtual implementation during the COVID-19 pandemic in the United States and worldwide. While this transition was necessary to prevent COVID-19’s spread, employment of virtualization for procedures formerly conducted face-to-face may have a lasting effect on how future clinical trials are performed. Virtual trials may be cheaper, more efficient, and may reach broader, more remote communities of participants. Despite these benefits, virtual clinical trials risk introducing additional barriers to vulnerable population subgroups including low-literacy, low-income and elderly patients. Our study represents an early investigation into how the switch from in-person clinical trial recruitment and study implementation to virtual during the COVID-19 pandemic may influence the demographic characteristics of participants who are recruited into RCTs. Given that virtual RCTs are more likely to rely on technology for recruitment, onboarding, training and retention, it is particularly important to ensure that these new procedures do not deter individuals from low-income communities with traditionally lower access to and proficiency with technology.[25]

We evaluated differences in clinical factors, demographic characteristics, and technical literacy between patients recruited in-person and virtually in the T2 Coach study. No statistically significant differences were found in gender, language spoken at home, education level, employment status, combined family income, marital status, age, and HbA1c at baseline. Birth location, race and ethnicity, insurance, and BMI were significantly different between the in-person and virtually recruited patient populations, although this was not due to a lack of representation of underserved participants. More patients recruited virtually were Hispanic and born in another country, and a higher proportion of patients recruited virtually were uninsured, without any changes in the recruitment focus, indicating that low-income participants were not overlooked. Some of these observed differences may be due in part to differences in the patient demographics of the participating FQHCs, since in-person recruitment was limited to 2 out of the 6 FQHCs that were included in the study. Consent rates during virtual recruitment were comparable to recruitment that occurred in-person.

In addition to historic under-representation in RCTs, virtual RCTs and studies that test mHealth interventions introduce new challenges to recruiting vulnerable patients due to higher demands for technology access and digital literacy. For example, despite widespread mobile device proliferation, age remains a barrier to smartphone use in personal healthcare.[26] Contrary to these concerns, the study found no significant differences between the in-person group and the virtual group in mobile device proficiency, a characteristic that is directly relevant to the new eligibility criteria and, as a result, the one at the highest risk for introducing selection bias.

At the same time, our study suggests that some adjustments to recruitment, onboarding, and retention protocols are needed when transitioning from in-person to virtual RCTs. Specifically, our findings suggest that recruiting participants from medically underserved populations for a virtual RCT evaluating an mHealth intervention may require greater time, educational and technical support resources compared to in-person recruitment. After switching to a virtual protocol, a longer amount of time was needed to complete virtual training and many patients needed more support (for example, patients may not have known how to share their screen on Zoom calls and required guidance from interventionists). Participants were allowed to ask as many questions as they wanted to, which extended the amount of time and resources required to complete intervention training but led to successful patient recruitment and enrollment from vulnerable communities. A more easily understandable verbal consent process, like the one our study implemented after virtualization, may be helpful in recruiting medically underserved patients compared to the lengthier, more complex written consent form that we used during in-person clinical trial implementation.

Our study contributes to the growing body of literature examining how informatics-based interventions may be evaluated amongst broad study populations.[27,28] Informatics interventions are widely impacting healthcare research and increasingly studied in RCTs.[29] Mobile health interventions have potential to improve patient health outcomes [30–32] at lower cost and scale but have yet to be widely studied in RCTs.[33,34] As mHealth and other informatics-based interventions are increasingly deployed in RCTs [35–37] and clinical practice, it is critical that they are conducted in a way that mitigates rather than exacerbates the historical under-representation of medically underserved patient populations that have characterized RCTs of the past.[10,38,39] Our study is an important contribution in showing how patients from medically underserved communities may be successfully recruited into a RCT for an mHealth intervention.

This work also expands on research into the recent proliferation of clinical trial virtualization catalyzed by the COVID-19 pandemic [40–42] and contributes to the limited body of literature on recruiting medically underserved populations for virtual clinical trials. [43] This study is the first to our knowledge to present patient recruitment amongst medically underserved patients for a virtual RCT that evaluates an informatics intervention. Our study builds on prior research that has shown the potential for informatics-based interventions to be successfully implemented amongst medically underserved populations [44] but that these patients often require higher training and support needs to use technologies,[45] which interventions may not account for. These findings underscore the need for investment into building intervention teams that account for higher subject training needs, the development of robust intervention training materials, and resources to expand technology access for vulnerable study participants. [46–49]

Our study also has limitations, including an age exclusion criterion (patients had to be 65 or younger), which prevented us from examining the impact of virtualization on older individuals. Our study has a relatively small sample size, which enabled interventionists to give patients individual attention, which may be difficult to scale for larger RCTs. Our study had a high number of foreign-born participants and was implemented in an urban setting in a large metropolitan area, which limits its generalizability. Finally, our study does not examine demographic, clinical, and technical literacy characteristics of our participants versus the communities they are from, which is particularly critical in the context of an mHealth intervention, which may reduce access due to their high technical literacy demands. These limitations raise important questions for future research, including how virtualized RCTs can recruit patients who are representative of their communities, and how patient engagement and self-management (such as home testing of blood glucose) affects the implementation and intervention effectiveness. Additionally, since our study’s scope is limited to baseline recruitment in a T2DM trial, future research can investigate how virtualization protocols impact patient retention and follow up.

Our findings suggest that transitioning to either hybrid or fully virtual procedures for clinical trials of informatics interventions will not necessarily exclude patients from medically underserved populations and can be considered as a plausible alternative or addition to the more expensive and labor-intensive in-person procedures. As awareness of issues surrounding inclusion and bias expands in the informatics community, [50,51] our study provides insights into ways to minimize barriers for historically excluded patients for participating in RCTs and interventions that use mHealth and other informatics-based solutions to improve patient engagement, self-management and health outcomes. Future research in this area should focus on clinical trial virtualization’s impact on patient-centeredness, and whether these changes will affect recruitment of special populations, such as older adults, patients with disabilities, and patients in rural areas. In conclusion, this work provides insights into how new methods of virtualization for recruiting and retaining diverse study populations, can be more inclusive of patients with low technical literacy, as well as identify what implementation strategies are needed to prevent increasing disparities and enhance health equity for those with limited digital proficiency.

## FUNDING STATEMENT

This work is funded by The National Institute of Diabetes and Digestive and Kidney Diseases (NIDDK) (Grant #1 R01 DK113189-01A1) & NLM T15 Training Grant T15LM007079.

## COMPETING INTERESTS

The authors have no competing interests to declare.

## CONTRIBUTORSHIP STATEMENT

EC, AS, GH, JT, and LM contributed to conceptualization. EC, TL, and AC contributed to data curation. EC conducted formal analysis. EC and HJ contributed to methodology. EC contributed to writing (original draft). EC, AC, TL, JC, AS, PD, HJ, GH, JT, and LM reviewed and edited the manuscript. AC, PD, and TL contributed to project administration. LM contributed to supervision and funding acquisition. All authors had final responsibility for the decision to submit for publication and agree to be accountable for all aspects of the work in ensuring that questions related to the accuracy or integrity of any part of the work are appropriately investigated and resolved.

## DATA AVAILABILITY STATEMENT

The data that support the findings of this study are available from the corresponding author upon reasonable request.

## ACKNOWLEDGMENTS

The study team would like to thank the FQHCs that participated in this study: Open Door Family Medical Center (Ossining, NY), Open Door Family Medical Center (Port Chester, NY), Metropolitan Family Health Network (Jersey City, NJ), Morris Heights Health Center (The Bronx, NY), Family Physician Health Center at NYU Langone (Brooklyn, NY), and Bedford Stuyvesant Family Health Center (Brooklyn, NY).

## Notes

### Competing Interest Statement

The authors have declared no competing interest.

### Author Declarations

In March 2020, the T2 Coach study team paused recruitment due to the COVID-19 pandemic, restrictions on in-person gatherings in New York City, and to ensure the safety of research and site staff, as well as the FQHC patients. During the spring and summer of 2020, the team developed a new set of protocols and strategies for implementing all components of the project virtually, including recruitment, onboarding, training, and all follow-up and retention activities, and secured IRB approvals for the new set of procedures. Institutional Review Boards at WCG Clinical, Columbia University Irving Medical Center, and Clinical Directors Network reviewed the T2 Coach Study and provided ethical approval.

